# Growth Efficiency of Donor Human Milk Compared With Maternal Milk and Formula in Preterm Infants

**DOI:** 10.1101/2025.10.03.25337279

**Authors:** Joseph H. Chou, Jonathan A. Berken, Brett D. Nelson, Juan D. Matute

## Abstract

**Importance:** Human milk is beneficial for preterm newborns, and pasteurized donor human milk (PDHM) is increasingly used when mother’s own milk (MM) is insufficient. However, PDHM has been associated with suboptimal growth outcomes.

**Objective:** To quantify the comparative clinical effectiveness of PDHM, MM, and formula in supporting daily weight gain and growth outcomes during the birth hospitalization.

**Design:** Retrospective cohort study of infants born before 34 weeks’ gestation between 2016 and 2024

**Setting:** Five newborn nurseries across the Mass General Brigham healthcare system (Boston, MA), including one Level IV, one Level III, and three Level II newborn units.

**Participants:** A total of 2,635 preterm infants born before 34 weeks’ gestation between 2016 and late 2024 were included in the daily weight gain analysis, contributing 40,007 days of eligible enteral intake data for evaluation of in-hospital growth outcomes. For the analysis examining the association between PDHM use and discharge weight, 2,719 infants met eligibility criteria.

**Exposures:** PDHM, MM, formula

**Main Outcomes and Measures:** The primary outcome was daily weight gain (g/kg/day). The secondary outcome was the change in weight Z-score from birth to discharge.

**Results:** In adjusted analyses, an equal volume of PDHM was associated with 74% of the daily weight gain achieved with MM (p < 0.0001, 95% CI 70%–77%), and formula with 115% of MM (p < 0.0001, 95% CI 111%–119%). During the birth hospitalization, infants in the highest quartile of PDHM exposure had a significantly greater decline in weight-for-age Z-score compared with those with no exposure (–0.09, p = 0.005), while no differences were observed in the lower quartiles.

**Conclusions and Relevance:** Growth outcomes varied by PDHM exposure, with a significant decline observed only in the highest quartile, while lower exposure levels showed no significant effect. Formula-fed infants were observed to have greater weight gain than either PDHM or MM. These results highlight the need for close growth monitoring and timely nutritional adjustments in preterm infants, particularly when PDHM is the primary source of enteral nutrition.

**Question:** How does pasteurized donor human milk (PDHM) compare with mother’s own milk (MM) and formula in supporting in-hospital growth among preterm infants born at or before 34 weeks’ gestation?

**Findings:** In this multicenter retrospective cohort study involving > 2500 preterm infants contributing over 40,000 infant-days of enteral data, PDHM correlated with significantly reduced daily weight gain compared with MM, whereas formula supported greater weight gain than either human milk source. At discharge, infants with the highest quartile of PDHM exposure showed a greater reduction in weight-for-age Z-score than infants with no PDHM exposure. No association was observed in the lower quartiles.

**Meaning:** Our findings underscore the need for close growth monitoring and potentially greater nutritional supplementation in relation to PDHM exposure.

## Introduction

Human milk is widely recognized as the gold standard for enteral nutrition in preterm infants^1–3^. When mother’s own milk (MM) is unavailable or insufficient, pasteurized donor human milk (PDHM) serves as an alternative^1,2^. Both MM^4^ and PDHM^5^ have been shown to reduce the risk of necrotizing enterocolitis (NEC). However, PDHM has been associated with slower growth than MM or formula^5,6^ posing a nutritional challenge for the very low birth weight (VLBW) infant (< 1500g) who is at heightened risk of growth impairment^7–9^.

In recent years, the expansion of milk bank infrastructure and growing implementation of hospital-based PDHM programs have significantly augmented the availability and utilization of donor milk in neonatal intensive care units (NICU) across the United States^10,11^. PDHM is now increasingly used both as a supplement to MM and, in some cases, as the primary source of enteral nutrition^1,2^. This evolving practice has prompted closer examination of neonatal outcomes associated with donor milk exposure. In this regard, randomized controlled trials (RCT) have demonstrated that PDHM in preterm infants reduces the incidence of NEC, albeit with a trade-off in postnatal growth velocity^12,13^. The Donor Milk for Improved Neurodevelopmental Outcomes (DoMINO) trial, a double-blind RCT, showed that supplementation with PDHM significantly reduced the risk of NEC compared to formula but was associated with slower postnatal weight gain^12^. Similarly, the Mother’s Own Milk (MOM; Milk) trial, which compared PDHM with preterm formula in VLBW infants, found a lower incidence of NEC in the PDHM group, though slower weight gain remained a persistent issue^13^. While these RCTs have established broad associations between feeding modality and growth^12,13^, few investigations have provided high-resolution, feed-level quantification of these effects while controlling for confounding factors.

This study aimed to rigorously measure the effects of MM, PDHM, and formula on in-hospital growth trajectories in preterm infants. Leveraging multi-year, daily feed and weight data from over 2,500 infants across five hospitals, we examined patterns of PDHM utilization, assessed their impact on growth outcomes, and identified clinical contexts in which intensified longitudinal monitoring of weight gain may be warranted to balance optimal growth with NEC prevention. By harnessing granular nutritional data from a heterogeneous cohort, this study enables a nuanced and controlled analysis of how each feeding type independently contributes to both short- and longer-term growth. This level of precision offers novel, actionable insights to inform nutrition strategies tailored to the complex and evolving needs of this high-risk population.

## Methods

### Data Acquisition and Processing

Data were obtained from the Mass General Brigham (MGB) Enterprise Data Warehouse, which included comprehensive electronic health record information from five MGB-affiliated newborn nurseries (Boston, MA): one Level IV, one Level III, and three Level II. The dataset encompassed all newborns admitted to an MGB nursery, their gestational age (GA), birthweight (BW), sex assigned at birth, enteral nutrition, and growth parameters. Enteral feeding data were collected for all documented feeds, specifying the type of nutrition (PDHM, MM, formula), reported caloric density (kcal/ounce), and volume administered. Growth data included all recorded weights throughout hospitalization. Weight-for-age was calculated using the Fenton 2013 preterm growth charts, following previously established methods^14,15^.

Extreme outliers were excluded using the following criteria: weight measurements beyond ±4 standard deviations from the mean (i.e., below the 0.003 or above the 99.997 percentile), single enteral feed volumes exceeding 300 mL, and daily weight changes (g/kg) outside the 0.1–99.9 percentile range. These outliers likely resulted from data entry errors or clinical interventions, and their exclusion was essential to enhance the accuracy and reliability of growth assessments in preterm infants receiving various types of enteral nutrition.

### Eligibility Criteria

Two distinct cohorts were defined for separate analyses of neonatal growth outcomes: one for daily weight gain and another for discharge growth.

### Daily Weight Gain Analysis

Infants were eligible for inclusion in the day-to-day weight gain analysis if they met the following criteria: (1) postmenstrual age (PMA) ≤34 weeks at the time of measurement; (2) receipt of at least 100 mL/kg/day of enteral nutrition; and (3) availability of consecutive daily weight measurements, from which the outcome of g/kg/day weight change was calculated. The 34-week PMA threshold was chosen because breastfeeding is minimal or absent below this age, with a mean of only 0.02 breastfeeds per analysis day, many of which were non-nutritive, allowing for a clearer evaluation of enteral nutrition’s impact on growth. Beyond this age, substantial direct breastfeeding may occur, complicating accurate quantification of intake volumes.

### Birth Hospitalization Growth Outcomes Analysis

Infants were included in the discharge growth trajectory analysis if they met all of the following criteria: (1) GA at birth ≤34 weeks; (2) admission to an MGB Level II, III, or IV newborn nursery within the first two days of life; (3) survival to hospital discharge; (4) discharge weight ≥ BW, from which the outcome of change in weight Z-score from birth to discharge was calculated; (5) discharge between 35 and 50 weeks PMA; and (6) documented weight measurement within one day of discharge. Although rarely necessary, missing daily weight values were linearly interpolated to calculate daily enteral intake in mL/kg/day.

### Model Development and Statistical Analysis

To quantify the relationship between enteral nutrition and daily weight gain, we developed a multivariable linear regression model with intake volume in mL/kg/day of MM, PDHM, and formula and additional caloric fortification in kcal/kg/day as the primary predictors. In accordance with local practice, MM and PDHM were assumed to provide 20 kcal/oz prior to fortification, ensuring accurate estimation of the caloric contribution from added fortifiers. For formula feeds, additional fortification was defined as the extra kcal/kg/day above a baseline of 20 kcal/oz. Sex assigned at birth, PMA centered at 29 weeks (for convenience in interpretation, chosen to be near-median PMA of cohort), and current weight-forage Z-score were included as covariates consistent with established growth chart parameters^14,15^.

To assess the pairwise relative impact of each feed type, we compared model coefficients using Wald tests with an asymptotic chi-squared statistic, contrasting an unrestricted model with a linearly constrained model^16^. Confidence intervals for the ratios of coefficients were calculated using the delta method^17^. We accounted for within-infant correlations and institutional variability, by employing a linear mixed-effects model with random intercepts for individual infants and hospital sites. Year was included as a random effect to control for temporal changes in feeding practices and growth outcomes.

This study was approved by the Mass General Brigham Institutional Review Board.

## Results

### Daily weight gain with MM, PDHM, and Formula

A total of 2,635 preterm infants met eligibility criteria for inclusion in the daily weight gain prediction model. The cohort had a median GA at birth of 31 2/7 weeks, a median BW of 1,495 grams, and contributed a median of 11 days of weight measurements (IQR 5–23) **(Table 1)**, yielding a total of 40,007 days of enteral intake and growth data analyzed. The mean daily weight gain was 24 g/kg at a median chronological age of 18 days (IQR: 10–30), corresponding to a median PMA of 32 2/7 weeks. The average enteral feed volume was 150 mL/kg/day, comprising on average 28 mL/kg of PDHM, 107 mL/kg of MM, and 15 mL/kg of formula, along with an additional 25 kcal/kg/day from fortification **(Table 2)**. Daily exposure to high volumes of PDHM (≥150 mL/kg/day) became more frequent after 2020. From 2017–2019, only 0.9% of hospitalization-days (460 of 51,182) reached this threshold, compared with 3.2% (2,477 of 77,403) in 2020– 2024, representing a 3.6-fold increase (p < 0.0001).

**Table 1.** Baseline characteristics of 2,635 infants contributing enteral intake and growth data.

| Characteristic | N = 2,635<sup>†</sup> |
| --- | --- |
| Gestational age (weeks) | 31.29 (29.14, 32.57): range 22.71 - 33.71 |
| Birth weight (grams) | 1,495 (1,130, 1,835): range 410 - 3,310 |
| Birth weight Z-score | -0.07 (0.84) |
| Number of days analyzed | 11 (5, 23) |
| Sex |  |
| F | 1,245 (47%) |
| M | 1,390 (53%) |
| Birth year |  |
| 2016 | 157 (6.0%) |
| 2017 | 306 (12%) |
| 2018 | 296 (11%) |
| 2019 | 291 (11%) |
| 2020 | 281 (11%) |
| 2021 | 334 (13%) |
| 2022 | 330 (13%) |
| 2023 | 311 (12%) |
| 2024 | 329 (12%) |

<sup>†</sup> Median (Q1, Q3): range Min - Max; Mean (SD); Median (Q1, Q3); n (%)

**Table 2.** Enteral intake and growth data comprising 40,0007 infant-days.

| Characteristic | N = 40,007<sup>†</sup> |
| --- | --- |
| Daily weight change (g/kg/day) | 24 (34) |
| Weight (grams) | 1,476 (374) |
| Weight Z-score | -0.72 (0.66) |
| Age (days) | 18 (10, 30) |
| Postmenstrual age (weeks) | 32.29 (30.86, 33.29) |
| Total volume (mL/kg/day) | 150 (16) |
| PDHM (mL/kg/day) | 28 (54) |
| MM (mL/kg/day) | 107 (63) |
| Formula (mL/kg/day) | 15 (41) |
| Fortification (kcal/kg/day) | 25 (11) |

<sup>†</sup> Mean (SD); Median (Q1, Q3)

The multivariable linear regression model **(Table 3)** estimated daily weight gain (g/kg/day) as a function of enteral intake and clinical variables. Specifically, each additional 1 mL/kg/day of intake from PDHM, MM, and formula was associated with weight gain increases of 0.178, 0.242, and 0.279 g/kg/day, respectively (all p < 0.0001).

**Table 3.** Regression model of daily weight gain (g/kg/day) by enteral feed type, adjusted for volume and fortification.

| Characteristic | Beta | 95% CI | p-value |
| --- | --- | --- | --- |
| (Intercept) | -25.2 | -28.4, -22.0 | <0.001 |
| PDHM (mL/kg/day) | 0.18 | 0.15, 0.20 | <0.001 |
| MM (mL/kg/day) | 0.24 | 0.22, 0.26 | <0.001 |
| Formula (mL/kg/day) | 0.28 | 0.25, 0.30 | <0.001 |
| Fortification (kcal/kg/day) | 0.30 | 0.27, 0.33 | <0.001 |
| PMA relative to 29 weeks | 2.16 | 1.96, 2.36 | <0.001 |
| Current weight Z | 0.31 | -0.20, 0.82 | 0.2 |
| Sex |  |  |  |
| F | — | — |  |
| M | 1.55 | 0.89, 2.21 | <0.001 |

Abbreviation: CI = Confidence Interval

In adjusted analyses, an equal volume of PDHM was associated with 74% of the daily weight gain achieved with MM (p < 0.0001, 95% CI 70% - 77%), and formula with 115% of MM (p < 0.0001, 95% CI 111% to 119%).

Furthermore, every kcal/kg/day of fortification contributed to 0.30 g/kg/day of weight gain. Additional predictors significantly associated with increased weight gain included PMA, with each week beyond 29 weeks contributing 2.16 g/kg/day (p < 0.001), and male sex associated with a 1.55 g/kg/day greater gain (p < 0.001). Weight Z-score was not a significant predictor (p = 0.23. The intercept (–25.2 g/kg/day) represented the predicted weight change for a 29-week female infant at the 50th weight percentile receiving no enteral nutrition. Mixed-effects modeling revealed minimal variability across individual infants and hospital sites (SD < 0.001 g/kg/day), with birth year accounting for only modest variation (SD = 0.93 g/kg/day).

### Birth Hospitalization Growth Analysis

For the analysis of the association between PDHM use and longitudinal growth, 2,719 infants met eligibility criteria **(Table 4)**. The median GA at birth was 32 2/7 weeks, with a median BW of 1,670 g. The median length of stay was 38 days, and the median PMA at discharge was 37 4/7 weeks. Across the entire hospitalization, mean daily enteral intakes were 13 mL/kg/day of PDHM, 87 mL/kg/day of MM, and 35 mL/kg/day of formula. The mean change in weight Z-score from birth to discharge was −0.66.

**Table 4.** Cohort characteristics and feeding profiles of 2,719 infants included in the birth hospitalization growth analysis.

| Characteristic | N = 2,719<sup>†</sup> |
| --- | --- |
| Gestational age (weeks) | 32.29 (30.00, 33.43): range 22.71 - 34.00 |
| Birth weight (grams) | 1,670 (1,245, 1,995): range 410 - 3,565 |
| Birth weight Z-score | $-0.14$ (0.86) |
| Sex |  |
| F | 1,304 (48%) |
| M | 1,415 (52%) |
| Birth year |  |
| 2016 | 122 (4.5%) |
| 2017 | 294 (11%) |
| 2018 | 307 (11%) |
| 2019 | 318 (12%) |
| 2020 | 290 (11%) |
| 2021 | 353 (13%) |
| 2022 | 372 (14%) |
| 2023 | 336 (12%) |
| 2024 | 327 (12%) |
| Length of stay (days) | 38 (25, 63) |
| Discharge PMA (weeks) | 37.57 (36.43, 39.43) |
| Change in weight Z score | $-0.66$ (0.50) |
| PDHM (mL/kg/day) | 13 (18), 6 (1, 20): range 0 - 130 |
| MM (mL/kg/day) | 87 (49), 107 (43, 128): range 0 - 156 |
| Formula (mL/kg/day) | 35 (42), 14 (0, 65): range 0 - 161 |
| PDHM category (average daily mL/kg/day) |  |
| 0 | 382 (14%) |
| [0.0162,2] | 585 (22%) |
| (2,9.07] | 584 (21%) |
| (9.07,21.9] | 584 (21%) |
| (21.9,130] | 584 (21%) |

<sup>†</sup> Median (Q1, Q3): range Min - Max; Mean (SD); n (%); Median (Q1, Q3); Mean (SD), Median (Q1, Q3): range Min - Max

Mean daily volume of PDHM across the hospitalization increased steadily over the study period **(Figure 1 A)**. At the patient level, changes in weight Z-score from birth to discharge varied by quartile of PDHM exposure **(Table 5)**. Infants who were never exposed to PDHM had a mean ΔZ-score of −0.65. Compared to no exposure, low-to-moderate PDHM exposure did not differ significantly from the no-PDHM group: –0.05 in the lowest quartile (p = 0.15), +0.06 in the second quartile (p = 0.081), and +0.05 in the third quartile (p = 0.11). However, weight Z-score declined in the highest PDHM exposure quartile (–0.09, p = 0.005), suggesting a non-linear relationship between PDHM volume and growth **(Figure 1 B)**.

**Table 5.**
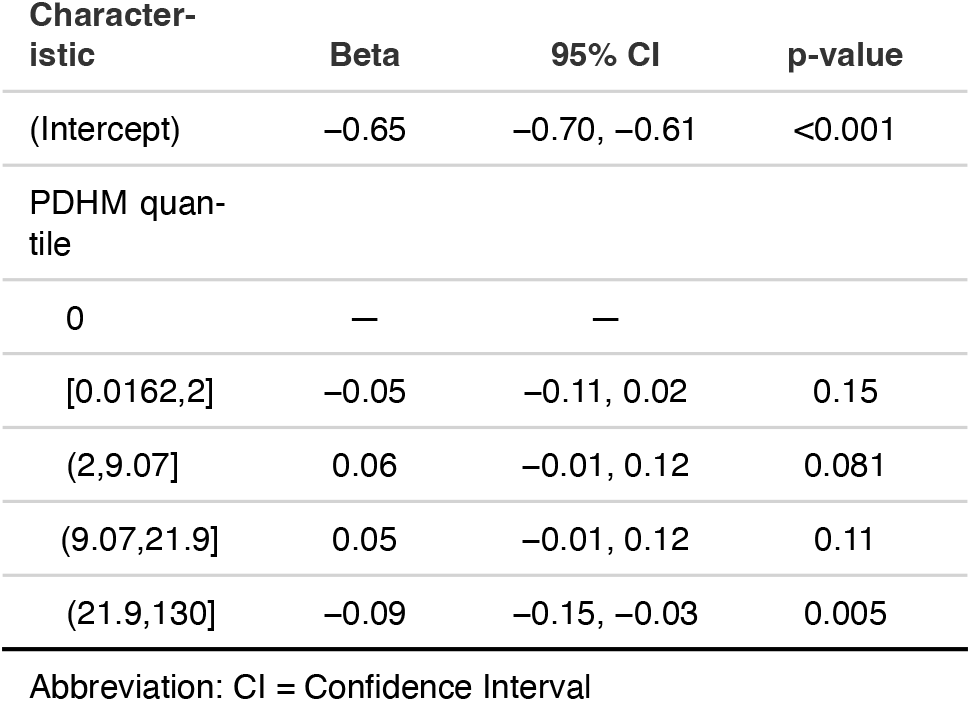
Association between PDHM intake and in-hospital growth.

| Characteristic | Beta | 95% CI | p-value |
| --- | --- | --- | --- |
| (Intercept) | -0.65 | -0.70, -0.61 | <0.001 |
| PDHM quantile |  |  |  |
| 0 | — | — |  |
| [0.0162,2] | -0.05 | -0.11, 0.02 | 0.15 |
| (2,9.07] | 0.06 | -0.01, 0.12 | 0.081 |
| (9.07,21.9] | 0.05 | -0.01, 0.12 | 0.11 |
| (21.9,130] | -0.09 | -0.15, -0.03 | 0.005 |

Abbreviation: CI = Confidence Interval

**Figure 1.**
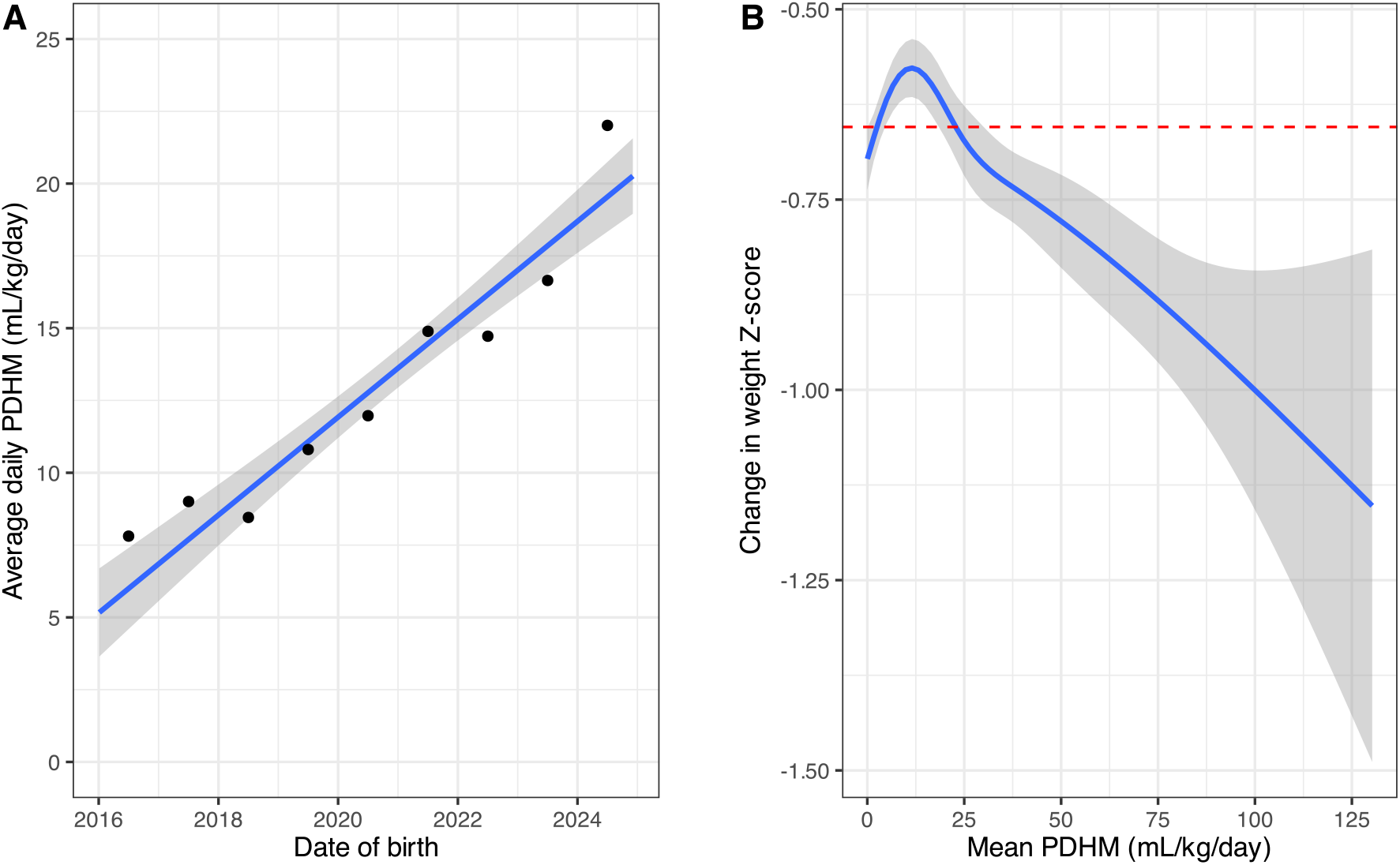
(A) Average daily PDHM intake during hospitalization by birth year and (B) relationship between PDHM intake and weight Z-Score change during birth Hospitalization. In (A), the blue line represents the linear regression trend across all individual patients, with the grey shaded area indicating the standard error of the mean. Black points denote the mean daily PDHM intake for all patients born in each respective year. In (B), the blue line shows the smoothed conditional mean change in weight Z-score (ΔZ) relative to PDHM intake, with the grey shaded area representing the standard error of the mean. The dashed red line indicates the mean ΔZ of –0.65 observed in infants with no PDHM exposure.

## Discussion

In this large, multicenter cohort of preterm infants born at or before 34 weeks’ GA, we observed a steady increase in PDHM use, consistent with trends reported elsewhere in the United States^10,11^. This pattern likely reflects its established role in NEC prevention^4,5^, the expanding availability of non-profit and commercial for-profit milk banks^11^, updated NICU policies and advocacy efforts prioritizing human milk for VLBW infants^2,3,18^, and broader governmental and private financial coverage^19^. However, this trend may also suggest increased reliance on PDHM as a prolonged feeding strategy rather than its original intended use as a short-term bridge to MM. Supporting this observation, daily intake of ≥150 mL/kg/day of PDHM increased 3.6-fold in preterm newborns born after 2020 compared with those born before.

Quantifying the independent effects of enteral nutrition type on postnatal weight gain, we found that formula supported the most daily growth, followed by MM, whereas PDHM was associated with the slowest rate of weight gain. We also observed a non-linear relationship between PDHM exposure and hospitalization growth outcomes. Compared with infants who received no PDHM, those in the low to moderate exposure range (i.e., first, second, and third quartiles of PDHM intake during hospitalization) showed a trend toward stable or slightly less negative weight Z-score changes. In contrast, the change in Z-score was significantly more negative at the highest quartile of PDHM exposure, compared with no PDHM exposure. These results indicate that modest PDHM use, likely clustered early in the hospital course reflecting its role as a bridge in most NICUs^1,5^, may be growth-neutral or mildly beneficial, while prolonged PDHM feeding is associated with reduced somatic growth.

Our findings of attenuated growth trajectories with increased use of PDHM reflect the inherent nutritional limitations of PDHM, including lower protein concentration^20–24^, loss of bioactive factors (e.g., stem cells, immunoglobulins, cytokines, enzymes) due to pasteurization^20,21,23–27^, and considerable variability in nutrient composition driven by donor characteristics^21,22^, lactation stage^28^, milk pooling practices^29^, and processing methods (e.g., Holder pasteurization versus high-temperature short-time pasteurization)^21,24,25,30^. This analysis demonstrates that the nutritional impact of feed type is not merely theoretical or compositional but clinically meaningful, producing measurable differences in growth outcomes. Therefore, current fortification practices likely do not fully meet the nutritional needs of infants receiving higher volumes of PDHM.

While this study provides compelling evidence on the relative effectiveness of PDHM in supporting daily growth, it also highlights key areas for future research. As a retrospective, observational study conducted within a single Boston-area hospital network, the ability to infer causality and generalize findings is inherently limited, and unmeasured factors such as illness severity, MM availability, maternal nutrition and overall health, or donor milk composition, may have influenced outcomes. In addition, important endpoints, including rates of NEC and long-term neurodevelopment, were not systematically captured. Building on these findings, a prospective RCT testing strategies such as enhanced caloric or protein fortification or tailored PDHM volumes would directly address growth challenges. This approach could provide definitive evidence to guide optimized, individualized, and safe feeding strategies in the NICU.

In conclusion, this study challenges the prevailing clinical assumption that equal volumes of PDHM, MM, and formula, after accounting for added caloric fortification, result in equivalent growth. By framing this assumption as our null hypothesis, we directly evaluated the real-world impact of current feeding practices. While PDHM remains a vital component of early neonatal nutrition and NEC prevention, it is substantially less effective in supporting growth compared to MM or formula. Shortterm PDHM use may be beneficial, but prolonged reliance or higher volumes of PDHM are associated with impaired somatic growth. Although our findings provide strong evidence that, volume-for-volume, PDHM is less capable of supporting daily growth compared with MM, overall growth across hospitalization is not necessarily affected. The primary conclusion of this study, that PDHM is a comparatively less effective substrate for promoting weight gain, remains unchanged. With vigilant clinical monitoring, differences in growth can be mitigated by adjusting PDHM volume or caloric fortification as needed. Whether such interventions might diminish PDHM’s established protective effects against NEC remains an open question. Future studies could investigate strategies to optimize growth while preserving these clinical benefits.

Our findings highlight the need for individualized, adaptive feeding strategies and potentially enhanced fortification protocols tailored to the fluctuating availability of MM and the nutritional requirements of PDHM-fed infants. Central to this approach is the early and sustained support of maternal lactation in the NICU, with attention to equity^31^. Taken together, this study demonstrates the relative effectiveness of PDHM in supporting daily growth and provides a hypothesis-generating exploration of strategies to mitigate its growth limitations while preserving its protective clinical benefits.

Clinical teams must recognize that feeding decisions are not a simple human milk-versus-formula choice but a dynamic process requiring continual assessment of nutritional adequacy, risks, and benefits throughout hospitalization. To further refine nutritional management, precise assessment of donor milk batches, including caloric content, macronutrient profiles, and distinctions between preterm and term PDHM, is critical for personalizing fortification and feeding plans. Finally, early or preemptive fortification strategies that account for the higher caloric requirements when using PDHM, particularly during transitions between feed types, are essential to meet the evolving nutritional needs of preterm infants and can be guided by our regression model. Future research should aim to define evidence-based nutritional approaches that balance safety, somatic growth, and long-term neurodevelopment in the preterm population. Innovations in alternative sterilization methods to preserve the bioactive properties of PDHM, real-time nutrient analysis, metabolomic profiling, and adaptive feeding algorithms responsive to the infant’s changing needs and available milk sources may hold promise in advancing care and improving outcomes. Ultimately, optimizing growth in preterm infants requires balancing the clear benefits of PDHM with proactive strategies to address its nutritional shortfalls through innovation and individualized care.

## Data Availability

All non-protected health information data produced in the present study are available upon reasonable request to the authors

## Acknowledgments

This work was supported by NIH grants K08DK133639 (JDM). The funders had no role in the study design, data collection, data analysis, data interpretation, or publication of this study.

## Author Contributions

- *Concept and design:* JHC, JAB, JDM
- *Acquisition, analysis, or interpretation of data:* JHC, JAB, BDN, JDM
- *Drafting of the manuscript:* JHC, JAB, JDM
- *Critical revision of the manuscript for important intellectual content:* JHC, JAB, BDN, JDM
- *Statistical analysis:* JHC
- *Supervision:* JHC, JDM

